# *In situ* transcriptional profile of a germinal center plasmablastic burst hints at *MYD88/CD79B* mutants-enriched Diffuse Large B-cell Lymphomas

**DOI:** 10.1101/2021.07.28.21261117

**Authors:** Vincenzo L’Imperio, Gaia Morello, Maria Carmela Vegliante, Valeria Cancila, Giorgio Bertolazzi, Saveria Mazzara, Beatrice Belmonte, Piera Balzarini, Lilia Corral, Arianna Di Napoli, Fabio Facchetti, Fabio Pagni, Claudio Tripodo

## Abstract

The germinal center (GC) reaction results in the selection of B-cells acquiring effector Ig secreting ability by progressing towards plasmablastic differentiation. This transition is associated with exclusion from the GC microenvironment. The aberrant expansion of plasmablastic elements within the GC fringes configures an atypical condition, the biological characteristics of which have not been defined yet. We investigated the *in situ* immunophenotypical and transcriptional characteristics of a non-clonal germinotropic expansion of plasmablastic elements (GEx) occurring in the tonsil of a young patient. Compared to neighboring GC and peri-follicular regions, the GEx showed a distinctive signature featuring key regulators of plasmacytic differentiation, cytokine signaling, and cell metabolism. The GEx signature was tested in the setting of diffuse large B-cell lymphoma (DLBCL) as a prototypical model of lymphomagenesis encompassing transformation at different stages of GC and post-GC functional differentiation. The signature outlined DLBCL clusters with different immune microenvironment composition and enrichment in genetic subtypes.

**Significance:** This report represents the first insight into the transcriptional features of a germinotropic plasmablastic burst, shedding light into the molecular hallmarks of B cells undergoing plasmablastic differentiation and aberrant expansion within the non-canonical setting of the GC microenvironment.

## Introduction

Within secondary lymphoid organs immune cells display topographic compartmentalization underlying functional commitment towards different stages of immune response induction and regulation. In lymphoid follicles, germinal centers (GCs) represent a complex specialized microenvironment sustaining B-cell proliferative bursts underlying somatic hypermutation and class-switch recombination of immunoglobulin (Ig) genes, communication with T cell subsets with helping function (Tfh), and interplay with specialized mesenchymal scaffolds (i.e. FDCs) (1). These events play through the dynamical iteration of elements between the dark (DZ), intermediate and light (LZ) zones of the GC (2), eventually resulting in the differentiation and displacement from the GC of cells acquiring effector capabilities through the synthesis and secretion of Igs (i.e. plasmablasts and plasma cells) (3). Alterations in the topographic compartmentalization of GC and extra-GC populations in lymphoid tissues are commonly observed in the setting of lymphoproliferative diseases, where the accumulation of cells with morphological or immunophenotypical features conflicting with their topographic localization represents a hallmark of histopathological analyses. This assumption reached its highest expression with the introduction of a diffuse large B cell lymphoma (DLBCL) prognostic sub-classification based on the presumed cell-of-origin (COO), as determined by gene expression profiling (GEP) (4-6).

We have investigated here an atypical germinotropic expansion of non-clonal, light-chain restricted B cells with plasmablastic features confined to a single enlarged GC structure in the tonsil of a young patient, through in situ immunolocalization analyses and high throughput digital spatial profiling. Comparing the features of the atypical germinotropic expansion (GEx) with those of topographically preserved DZ, LZ and peri-follicular (PERI) regions of interest (ROIs) we identified a unique transcriptomic profile of the GEx ROIs featuring the overexpression of transcripts involved in plasmacytoid differentiation, cytokine signaling, and cell metabolism.

To probe the reflection of the identified transcriptional signature in a setting of B-cell lymphomatous transformation embracing the full spectrum of GC- and post-GC differentiation, the discriminative 20 genes were used to cluster a large cohort of diffuse large B-cell lymphoma (DLBCL) transcriptomic data. The GEx signature highlighted two clusters with different overall survival in DLBCL, where cases with the highest expression of GEx hallmark genes were characterized by poorer prognosis.

## Materials and methods

### Clinical setting

This study started from the incidental finding of the reactive germinotropic plasmablastic expansion described in the Results section, in the tonsil of a young patient who underwent tonsillectomy for clinical hypertrophy. Sample was obtained and handled according to the Declaration of Helsinki. Informed consent for surgery and histopathological studies was obtained from the legal representatives. The case was included in the study 05/2018 approved by the University of Palermo Institutional Review Board.

### Histological, immunohistochemical and molecular analyses

Tonsillar tissue has been formalin-fixed and paraffin-embedded (FFPE) and 3 µm thick sections have been stained with hematoxylin and eosin (H&E). IHC has been performed at the Pathology Department of ASST Monza, San Gerardo Hospital, Monza, Italy using a Dako Omnis platform (Dako, Denmark) using antibodies directed against CD20 (L26), CD3 (Polyclonal), Bcl2 (124), Bcl6 (PG-B6p), CD21, ki-67 (Mib-1), IRF4 (MUM1), CD10, CD30 (Ber-H2), CD138 (Mi15), HHV8 (13B10), kappa and lambda light chains. Double immunohistochemistry has been performed for MUM1 and CD10 using 3-3’-diaminobenzidine (DAB) and 3-amino-9-ethylcarbazole (AEC) as chromogens, respectively. In situ fluorescence hybridization (FISH) study has been performed using a *IRF4/DUSP22* (6p25) Break Apart kit (Kreatech, Leica Biosystem, Germany) at the Pathology Department of Spedali Riuniti di Brescia. Quantitative evaluation of immunophenotypical markers was performed by applying the HALO image analysis software (v3.2.1851.229, Indica Labs) to regions selected on whole slide digital scans acquired using an Aperio CS2 slide scanner with the ImageScope software (v12.3.28013, Leica Biosystems, Germany).

Quantitative polymerase chain reaction (Q-PCR) to detect clonal immunoglobulin genes rearrangement was performed after laser microdissection on H&E-stained slides, using an LMD6 platform (Leica Microsystems, Germany).

### Digital spatial profiling

The transcriptional landscape of 15 different spatially-resolved regions of interests (ROIs) of the tonsil (5 peri/inter-follicular ROIs, 5 DZ and 5 LZ ROIs from morphologically normal follicles) and 9 ROIs from the GEx was determined by Digital Spatial Profiling on slides stained with CD271/NGFR (as an FDC marker to highlight the LZ) and CD20 (as a B-cell marker). The 24 selected and segmented ROIs were profiled using a GeoMx Digital Spatial Profiler (DSP) (NanoString, Seattle WA) as previously described (7), applying the Cancer Transcriptome Atlas panel (https://www.nanostring.com/products/geomx-digital-spatial-profiler/geomx-rna-assays/geomx-cancer-transcriptome-atlas/) (Supplementary Table 1).

### Bioinformatic Data Analysis

After quality check step, raw counts were normalized against the 75th percentile of signal from their own ROI and normalized data were used to perform PCA using FactoMine R package. For hierarchical clustering analysis of the ROIs the Euclidean distance metric across samples was considered and complete aggregation method was used for building tree within the R package hclust.

Differential expression analyses were carried out by applying the moderated t-test using the limma package (8); pairwise comparisons between GEx and DZ/LZ/Peri ROIs were considered. Upregulated/downregulated genes were selected for subsequent analysis if their expression values were found to exceed the threshold of 0.05 FWER (Bonferroni correction). The spatial GEx signature was assessed in the following Schmitz et al. dataset (6) and the clinical information was downloaded from the Schmitz et al. supplementary material.

After centering and scaling expression value, unsupervised hierarchical clustering analysis based on the GEx signature was perform to identify potential discriminative clusters based on Ward.D2 method on the Euclidean distance Survival analysis was performed using log-rank test implemented in “survival” R package. Differences in patient characteristics among groups were analyzed with the Fisher’s exact test.

CIBERSORTx (http://cibersortx.stanford.edu) (9) has been used to calculate proportions of microenvironment cell included in the LM22 signature on Schmitz *et al*. (6) RNA-seq data of bulk tissues. This dataset was downloaded and analyzed using the authors’ normalization setting which included fragments per kilobase of transcript per million (FPKM) space. Bulk-mode batch correction (B-mode) was applied to mixture samples before imputing cell fractions and 1000 permutations were set for significance. Moreover, the SpatialDecon algorithm (10) was additionally used for Nanostring data (safeTME profile matrix was applied). The differences in CIBERSORTx cell fractions among clusters were investigated using the Wilcoxon-Mann-Whitney test. Similarly, the differences in SpatialDecon cell fractions among ROI subgroups were investigated using the Kruskal-Wallis test.

All statistical analyses have been performed using R statistical software (v4.0.2, http://www.R-project.org).

### Data Availability

Normalized gene expression data generated in the Digital Spatial Profiling experiment are available in Supplementary Table 1

## Results

### Pathology of the tonsil

Histopathological analysis of the left tonsil from a young patient with clinical bilateral hypertrophy revealed, in a background of lymphoid follicles with hyperplastic features and preserved GC DZ, LZ and mantles, an isolated abnormal follicle with flattened mantle zone, an enlarged GC without evident DZ/LZ polarization, preservation of rare tingible body macrophages, and populated by a predominance of monomorphic plasmacytoid cells with immature morphology (Figure 1A, inset). Quantitative immunophenotypical characterization of reactive follicular (DZ and LZ) and peri-follicular (Peri) regions and of the atypical germinotropic plasmablastic expansion (GEx), highlighted conspicuous differences in the immune profile (Figure 1B-C). The reactive preserved GC DZ and LZ regions were characterized by B cells with strong CD20 expression, dense Ki-67 immunoreactivity (higher in the DZ), negativity for IRF4, except for scattered cells in a background of CD10-expressing cells, Bcl-2 negativity, slight T-cell infiltration (denser in the LZ), and no evidence of light chain restriction (Figure 1B-C). At contrast, the composition of the GEx displayed a CD20+ B cell phenotype, high Ki-67+ proliferative fraction, diffuse IRF4 positivity with IRF4+ cells co-expressing CD10, negativity for Bcl-2, and immunophenotypical restriction for lambda light chain (Figure 1B-C). Most of the cells populating the GEx also expressed CD138 (in the absence of CD30), indicating partial acquisition of a plasmablastic phenotype. Immunoistochemistry for HHV8 and *in situ* hybridization for EBER (EBV) proved negative (Supplementary Figure 1). On the basis of the GEx lambda light chain restriction, analysis of the Ig light and heavy genes rearrangement was performed on DNA extracted by laser microdissection of the GEx, which revealed a polyclonal profile (Supplementary Figure 2). The strong and diffuse immunoreactivity of IRF4 and the co-occurrence of IRF4/CD10 double-expressing elements prompted the analysis of *IRF4* gene rearrangement by fluorescence in situ hybridization (FISH), which did not reveal any abnormality (Supplementary Figure 3), allowing to exclude an *IRF4*-rearranged lymphoma.

**Figure 1.**
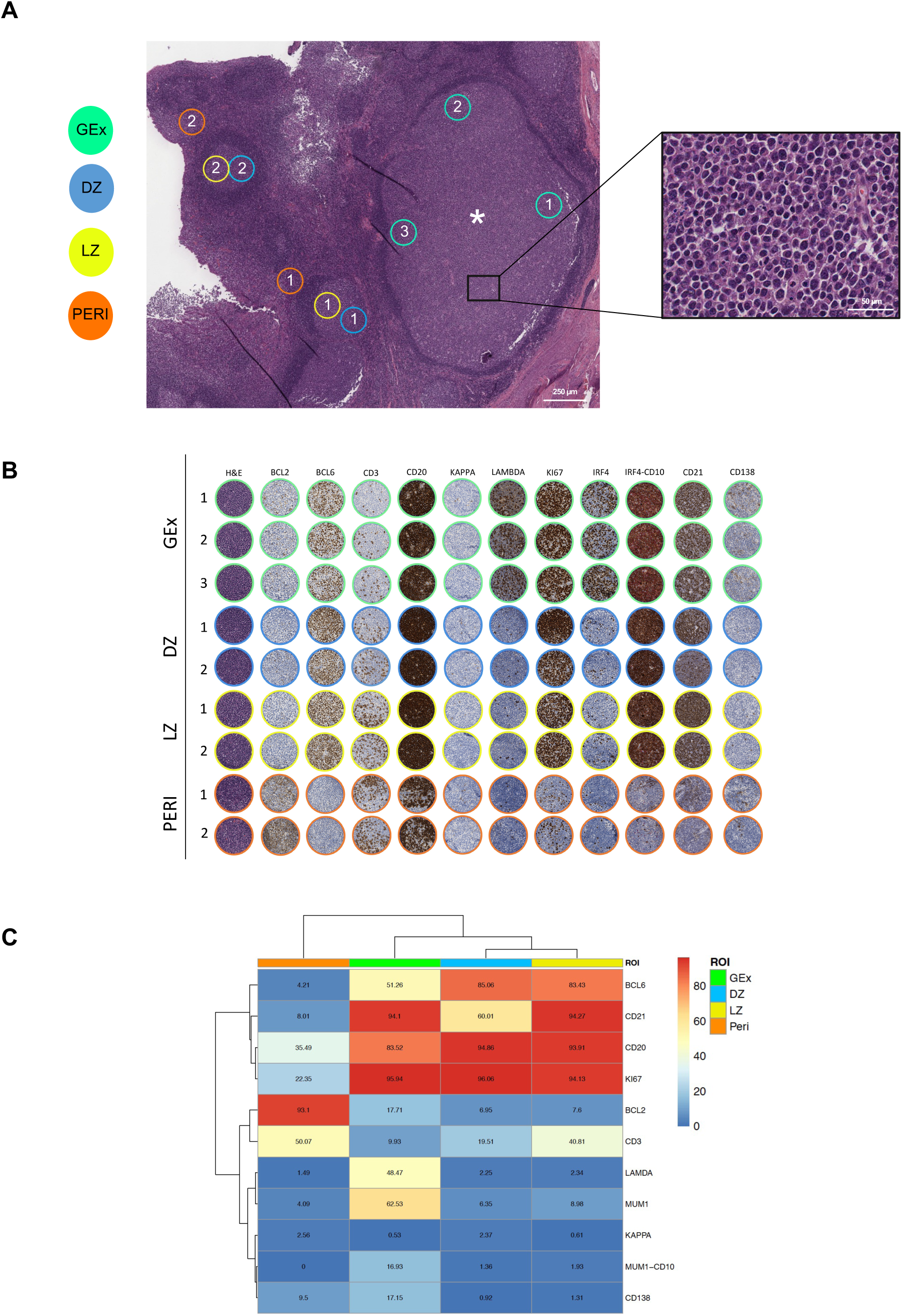
**A**, Digitalized slide selection of the Haematoxylin and Eosin (H&E)-stained section of the tonsil highlighting the presence of an aberrantly expanded germinal center (asterisk) characterized by the presence of elements with plasmacytoid morphology (inset). On the H&E, representative regions relative to germinal center dark zone (DZ) and light zone (LZ) areas, peri-follicular (Peri) areas, and germinotropic plasmablastic expansion (GEx) areas, are highlighted. Original magnification x50. **B**, Comparative analysis of H&E and IHC for Bcl-2, Bcl-6, CD3, CD20, Kappa and Lambda light chain, Ki67, IRF4, IRF4/CD10, CD2, CD138 in the DZ, LZ, Peri and GEx areas highlighted in A. **C**, Heatmap of the average expression of the quantitative immunohistochemical analysis of the markers evaluated in the DZ, LZ, Peri, and GEx areas highlighted in B.

### Digital Spatial Profiling of the GEx regions reveals a distinctive profile

We subsequently investigated the in situ transcriptional profile of the GC plasmablastic burst through the Nanostring GeoMx Digital Spatial Profiling technology. The expression of 1824 genes from key cancer-associated transcriptional programs (Supplementary Table 1) was determined on 5 DZ, 5 LZ and 5 PERI ROIs selected from morphologically/phenotypically preserved follicles/perifollicular areas, and on 9 GEx ROIs. We then asked whether GEx ROIs could be defined by a specific gene signature; to this aim, principal component analysis (PCA) and unsupervised hierarchical clustering were investigated. PCA revealed that ROIs segregated according to their spatial classification, with PERI regions showing neatly separated profiles from GC ROIs including DZ, LZ and GEx, which clustered together with other GC regions, showing some degree of intermixing with LZ ROIs (Figure 2A). Consistently, clustering analysis confirmed the same degree of relationship between the different ROIs (Figure 2B). mRNA expression of the transcripts relative to the IHC markers evaluated for quantitative immunopehnotypical analyses showed consistency with the protein expression pattern (Supplementary Figure 4). Pairwise differential expression analysis performed on the different ROIs allowed to identify candidate genes reflecting the distinctive profiles between the GEx ROIs in comparison with DZ LZ and PERI ROIs (Figure 2C-F). Among the 20 differentially expressed genes, 17 were significantly upregulated in GEx ROIs, while 3 were downmodulated (Figure 2C-F). GEx hallmark genes included, along with the plasma cell differentiation markers *PRDM1, IRF4, TNFRSF17* (BCMA) and *CD9*, genes involved in 2-oxoglutarate metabolism (*GOT2, IDH2)*, in IL17 pathway (*IL17RB, HSP90B1*) and cytokine signaling (*RASAL1, LTB*), in PI3K-Akt pathway (*SGK1, BCL2L1*), in lymphocyte activation (*ADA, SCL7A5, FCRL2*) and cell surface regulation of immune activation (*CD24, LILRB1*), in cell adhesion (*ANKRD28*) and response to abiotic (i.e. osmotic) stress (*SLK1*). Moreover, the long non-coding RNA *FAM30* was also listed among the GEx hallmarks.

**Figure 2.**
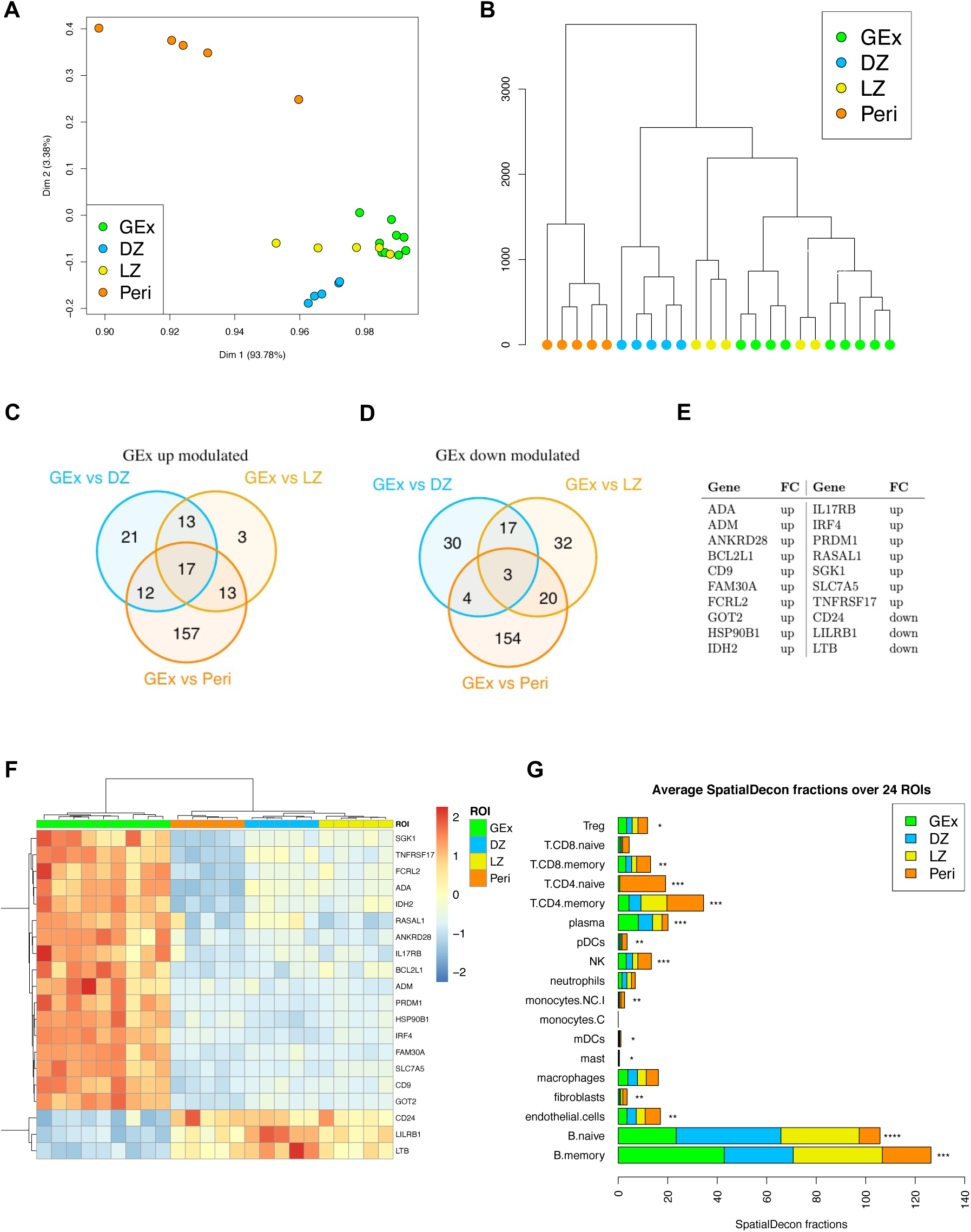
**A**, Two-dimensional principal component reduction of the DZ (n=5), LZ (n=5), Peri (n=5), and GEx (n=9) regions of interest (ROIs) profiles according to Digital Spatial Profiling of 1824 genes. **B**, Unsupervised hierarchical clustering of the 24 ROIs. **C**, Venn diagram of UP-modulated genes from three different comparisons (i.e., GEx vs DZ, GEx vs LZ, and GEx vs Peri). **D**, Venn diagram of DOWN modulated genes from three different comparisons (i.e., GEx vs DZ, GEx vs LZ, and GEx vs Peri). **E**, GEx signature genes. These genes are significantly differentially expressed in GEx in each comparison. **F**, Heatmap of differentially expressed genes in GEx as compared to DZ, LZ and Peri ROIs. The GEx signature shows a high discriminatory capacity between GEx ROIs and the other regions. **G**, Average SpatialDecon fractions of cell types in the four ROI subgroups. Kruskal-Wallis test has been applied to compare faction distributions among groups (Supplementary table 2).

### Spatial immune deconvolution of GEx ROIs shows enrichment in memory B cells and Plasma cells

Based on the evidence of a distinct transcriptional profile of GEx regions in comparison with other GC and perifollicular regions, we investigated the immune composition of the ROIs according to transcriptional deconvolution. A SpatialDecon (10) approach using the safeTME matrix was adopted, which highlighted that GEx ROIs displayed a different microenvironment composition as compared with canonical LZ and DZ GC ROIs, also differing from peri-follicular ROIs (Figure 3, Supplementary Table 2), further indicating that a perturbation of the normal GC milieu. Specifically, GEx ROIs were positively enriched in Plasma cells and memory B cells as compared with other ROIs (Kruskal-Wallis p-values < 0.001), while being poorly infiltrated by T cells (Figure 2G).

**Figure 3.**
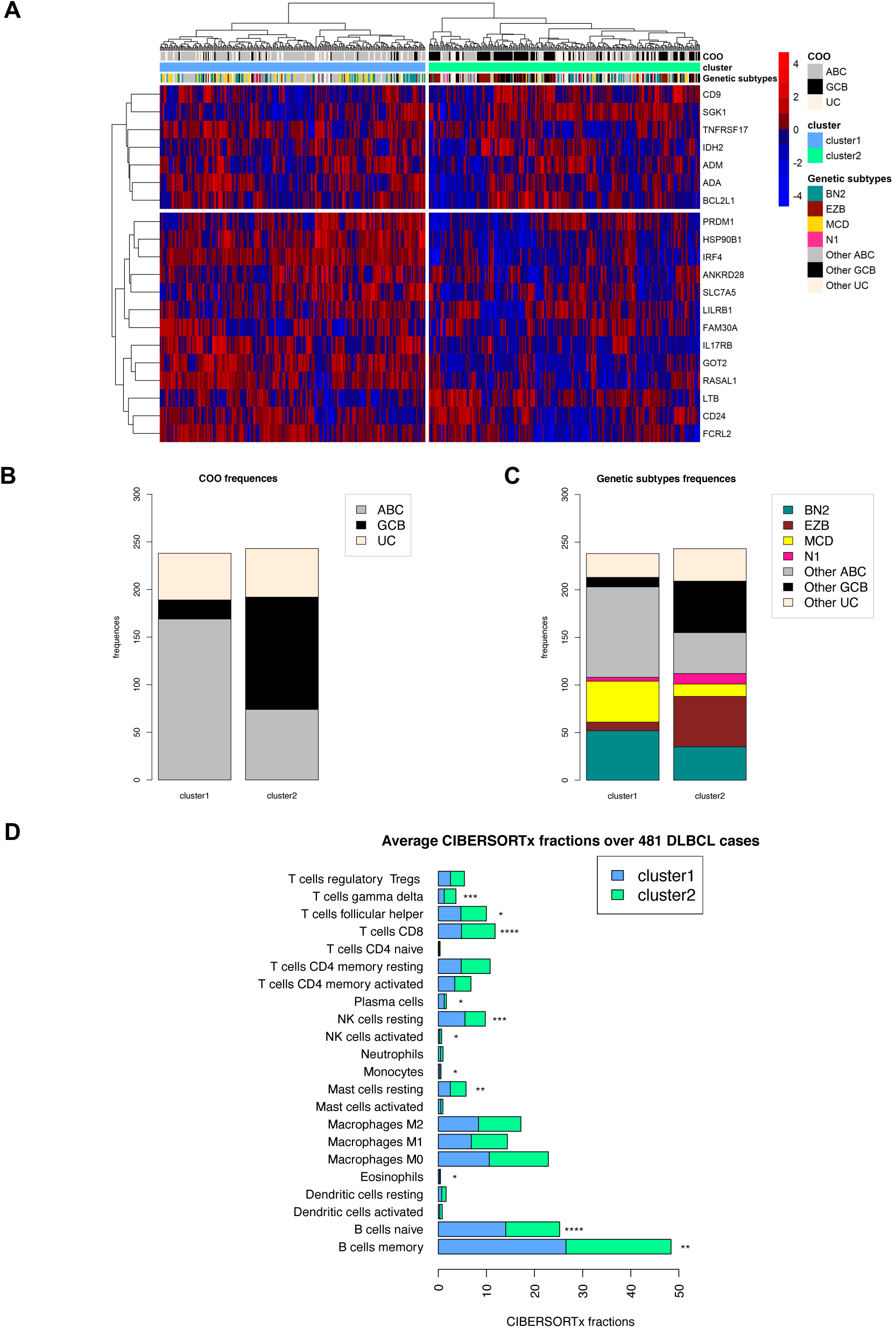
**A**, Unsupervised clustering analysis of the 481 DLBCL cases based on the GEx signature. It identifies two distinct clusters; the blue one is characterized by a higher gene expression, while a lower gene expression characterizes the green one. **B**, Barplot of cell of origin (COO) absolute frequencies observed among clusters. Cluster 1 is enriched by ABC cases, whereas cluster 2 is enriched by GCB cases (Fisher p-values < 10e-15). **C**, Barplot of genetic subtypes absolute frequencies observed among clusters (Fisher p-values reported in Supplementary Table 3). **D**, Average CIBERSORTx fractions of cell types in the four ROI subgroups. Wilcoxion-Mann-Withney test has been applied to compare faction distributions among groups (Supplementary Table 4).

### The GEx signature outlines DLBCL clusters with different enrichment in genetic subtypes and microenvironment composition

The molecular profiles of non-malignant GC compartments can be exploited to probe GC microenvironment imprints in B-cell lymphomas with different degree of relationship with GC subpopulations, such as DLBCL, in which the COO has shown prognostic significance in the setting of standard chemo-immunotherapy regimens (7). To investigate whether the transcriptional hallmarks identified in the GEx ROIs could be traced in the heterogeneous spectrum of B-cell malignant transformation recapitulated by DLBCL, we applied the GEx gene signature to a dataset of 481 DLBCL cases relative to Schmitz et al. (6). Based on the expression of the 20 genes of the GEx signature, DLBCL clustered into two main groups (Figure 3A), with the cluster 1 characterized by the overexpression of 17 genes (Supplementary Figure 5A) and by a trend towards a worse prognosis (Supplementary Figure 5B-C). We subsequently investigated the distribution of the major DLBCL genetic subtypes according to Schmitz and Colleagues (6) and found that the relative frequency of the subtypes was significantly different in the two clusters identified by the GEx signature (Figure 3B-C, Supplementary Tables 3 and 4). Specifically, cluster 1, which was characterized by the general overexpression of hallmark genes of the GEx ROIs, showed a neat enrichment in cases with *MYD88* and *CD79B* mutation co-occurrence (MCD, Fisher p-value<10e-05), a higher frequency of cases with BCL6 fusions and NOTCH2 mutations (BN2, Fisher p-value=0,02), and a markedly lower frequency of cases with EZH2 and BCL2 lesions (EZB, Fisher p-value<10e-9) (Figure 3C, Supplementary Tables 4) suggesting that the genes positively characterizing GEx ROIs underlie a specific biology related with MCD genetics, known to be enriched in ABC clones undergoing plasmablastic/plasmacytic commitment (6).

Prompted by the finding of a different microenvironment composition of GEx ROIs in comparison with other GC and peri-follicular ROIs, we estimated the microenvironment of the DLBCL clusters identified according to GEx signature, through CIBERSORTx deconvolution. Consistently with the deconvolution of in situ transcriptional profiles of GEx ROIs, cluster 1, which was characterized by up-regulation of GEx hallmark genes, showed a significant enrichment in memory B cells and plasma cells (Mann-Whitney p-values < 0.001, Supplementary Table 5), and a decrease in CD8 T cells and follicular T helper cells (Figure 3D), indicating that a link between the GEx that is a GC-related lsion, and lymphomatous clones enriched in specific non-GC genetics, cell-of-origin and microenvironment, may exist.

## Discussion

The transition of B cells undergoing selection and refinement of their IG receptor in the GC reaction towards effectors capable of Ig secretion implies the acquisition of plasmablastic/plasmacytoid features within the GC microenvironment. The spatial localization of these functional and phenotypical intermediates is still poorly characterized and depends on the dynamical modulation of chemotactic receptor/ligand axes interweaving with BCR-controlled programs (11). Proliferating cells with plasmablastic/plasmacytoid features accumulating within the GC therefore represents an element of atypia even in the setting of non-clonal events, and little is known about the *molecular signature* characterizing their transient state (12). In this report we phenotypically and transcriptionally characterized an immunoglobulin light chain restricted non-clonal atypical germinotropic expansion of plasmablastic cells, investigating differential features emerging from the comparison with neighboring GC and extra-GC regions. The GEx ROIs were characterized by the unique co-occurrence of IRF4 and CD10 expression, which highlighted a transitory state engendered by IRF4 control of GC exit (13) and CD10 ectopeptidase retention that can be observed in DLBCL with plasmablastic differentiation (14). On digital spatial profiling, a set of 20 genes were found differentially expressed in GEx ROIs as compared with neighboring DZ, LZ and PERI ROIs. The discriminating signature resulted positively enriched in the key transcription factors driving plasma cell differentiation IRF4 and PRDM1, and included the B-cell differentiation receptor BCMA involved in the transduction of trophic signals from APRIL and BAFF tumor necrosis factor superfamily ligands (15). Such molecular features supportive of a plasmablastic phenotype were also supported by the downregulation of CD24, a signal transducer negatively modulated in response to BCR activation and along plasmablastic transition (16). The GC localization of the plasmablastic expansion found resonance in the overexpression of CD9. The tetraspanin CD9 has been reported to mark a subset of B cells in the human GC characterized by plasmablastic differentiation and Blimp1 (*PRDM1*) expression (17). These CD9+ GC B cells more efficiently give rise to CD20-CD38+ plasmablasts as compared with their CD9-counterpart. Moreover, in the murine setting, the efficient plasmablastic/plasmacytic differentiation of CD9+ B cells is shared by non-GC B-cells endowed with prompt commitment to Ig-secreting effectors, such as B1 B cells and marginal zone B-cells subsets (18). By applying the GEx differentially expressed genes signature to a well-characterized (both transcriptionally and genetically) DLBCL dataset, we aimed at investigating whether the atypical status of non-clonal germinotropic plasmablastic expansion could be represented in the transcriptional signature of a subset of DLBCL of either ABC or GCB COO. Previous reports described a subset of ABC-DLBCL expressing *PRDM1*/BLIMP1 and demonstrated that loss of function of this antigen is harbinger of a poor prognosis (19). Expression of IRF4/MUM1 is routinely employed in the diagnostic setting for the distinction of non-GC subtypes based on immunohistochemical algorithms (20). Moreover, specific subtypes of large B-cell lymphomas characterized by IRF4 rearrangement have been recently described and recognized as independent entities in the most recent WHO classification (21). We report additional molecular markers potentially associated with plasmablastic commitment in the GC, including the receptor of IL17B/IL25 IL17RB, the overexpression of which marks lymphoplasmacytic lymphomas with mutant *MYD88*^L265P^ and *CXCR4*^WT^ (22). In the GC setting, IL17B/IL25 signaling could enforce NF-κB activity (23) through TRAF6, which cooperation with CD40 signaling is required for B-cell affinity maturation and plasma cell differentiation (24). From the genes positively and negatively characterizing GEx ROIs, no relevant clues emerge about the mechanisms leading to the atypical GC retention and expansion of the plasmablastic elements. Under normal conditions, the suppression of Bach2 and Pax5 transcripts, along with the down-regulation of Bcl-6, IRF8 and PU-1 and the activation of BLIMP1 and MUM1/IRF4, drive the development of PCs resulting from the GC reaction (1). Once their effector/memory fate is established, B cells escape from the GCs through the suppression of the BCL-6-induced “confinement factor” S1PR2 and the expression of pro-migratory receptors that are likely to be involved in GC exit, such as EBI2 and S1PR1 (25). The downmodulation of lymphotoxin beta transcript emerging from the GEx ROIs profiling can imply an impaired activation of the FDC meshwork by resident elements (26) which would in turn impact on the maintenance of a functional GC microenvironment licensing atypical plasmablastic expansion within the GC contexture. Indeed, some cases of abrupt/florid follicular hyperplasia have been described in which activated B cells are mainly localized in the GCs, partly twisting the normal follicular architecture and even showing immunohistochemical light chain (oligoclonal) restriction, leading to diagnostic concern for neoplasia (27). The perturbation of normal GC dynamics in GEx was also indicated by an overall depletion in the Th compartment on transcriptional deconvolution analysis, which was similar to that of the DZ, conflicting with the memory B and plasma cell enrichment. In DLBCL, GEx hallmark genes identified a subgroup with positive enrichment in MCD genetics and ABC COO, indicating that GC-related proliferations of plasmablastic elements may be transcriptionally linked with non-GC DLBCL. These cases characterized by a dyscrasia between cyto-architectural, phenotypical and topographic profile of a B-cell expansion with plasmablastic/plasmacytic features may help in finding a link with the pathogenesis specific DLBCL subsets (28) and reconcile an ABC-COO with GC-related lesions (29).

## Data Availability

Normalized gene expression data generated in the Digital Spatial Profiling experiment will be made available on request.

## Acknowledgements

The Authors wish to acknowledge Prof. Maurilio Ponzoni for helpful discussion. This study has been supported by the Italian Foundation for Cancer Research (AIRC) through the IG-2018 22145 Investigator Grant to C.T.; 5×1000 22759 Grant to C.T.; and by the Italian Ministry of Education, University and Research (MIUR) grant 2017K7FSYB to C.T.

